# Comparison of vaccine effectiveness against the Omicron (B.1.1.529) variant in patients receiving haemodialysis

**DOI:** 10.1101/2022.01.25.22269804

**Authors:** Katrina Spensley, Sarah Gleeson, Paul Martin, Tina Thomson, Candice L. Clarke, Graham Pickard, David Thomas, Stephen P. McAdoo, Paul Randell, Peter Kelleher, Rachna Bedi, Liz Lightstone, Maria Prendecki, Michelle Willicombe

## Abstract

**Background:** Emerging data suggest a reduction in SARS-CoV-2 vaccine effectiveness against Omicron SARS-CoV-2 infection. There is also evidence to show that Omicron is less pathogenic than previous variants. For clinically vulnerable populations, a less pathogenic variant may still have significant impact on morbidity and mortality. Herein we assess the clinical impact of Omicron infection, and vaccine effectiveness, in an in-centre haemodialysis (IC-HD) population.

**Methods:** One thousand, one hundred and twenty-one IC-HD patients were included in the analysis, all patients underwent weekly screening for SARS-CoV-2 infection via RT-PCR testing between 1^st^ December 2021 and 16^th^ January 2022. Screening for infection via weekly RT-PCR testing and 3-monthly serological assessment started prior to the vaccine roll out in 2020.

**Results:** Omicron infection was diagnosed in 145/1121 (12.9%) patients over the study period, equating to an infection rate of 3.1 per 1000 patient days. Vaccine effectiveness (VE) against Omicron infection in patients who had received a booster vaccine was 58 (23-75)%, p=0.002; VE was seen in patients who received either ChAdOx1, VE of 47(2-70)%, p=0.034, or BNT162b2, VE of 66 (36-81)%, p=0.0005, as their first two doses. No protection against infection was seen in patients who were partially vaccinated (2-doses), p=0.83. Prior infection was associated with reduced likelihood of Omicron infection, HR 0.69 (0.50-0.96), p=0.0289. Four (2.8%) patients died within 28 days of infection diagnosis, with no excess mortality was seen in patients with infection.

**Conclusion:** 3-doses of SARS-CoV-2 vaccines are required in ICHD to provide protection against Omicron infection.

---

Since its first detection in November 2021, the SARS-CoV-2 B.1.1.529 (Omicron) variant has rapidly became the dominant variant worldwide. Increased transmissibility and its ability to evade both vaccine and infection induced neutralising antibodies was of significant global concern, and was the impetus for the UK to expediate it’s booster vaccination programme^1^. In-vitro and animal models suggested that whilst demonstrating the unfavourable characteristics described, the pathogenicity of the Omicron variant was reduced^2,3^. The emerging real-world data appear to mirror the laboratory findings, with data showing a reduction in vaccine effectiveness against infection with Omicron but enhanced efficacy against severe infection requiring hospitalisation^4^.

For clinically vulnerable populations, a less pathogenic variant may still have significant impact on morbidity and mortality. People with end stage kidney disease (ESKD) receiving in centre haemodialysis (IC-HD) are one such patient group. Patients receiving IC-HD have attenuated responses to SARS-CoV-2 vaccines, with recent data suggesting patients who have received 3-doses of a heterologous vaccination regimen may have inadequate neutralising ability against Omicron, leaving them at significant risk of infection^5,6^. Herein we assess the clinical impact of Omicron infection, and vaccine effectiveness, in an IC-HD population followed up within a prospective longitudinal surveillance study at Imperial College London (HRA REC reference: 20/WA/0123).

## Results

One thousand, one hundred and twenty-one IC-HD patients were included in the analysis, all patients underwent weekly screening for SARS-CoV-2 infection via RT-PCR testing. Screening for infection via weekly RT-PCR testing and 3-monthly serological assessment started prior to the vaccine roll out in 2020 (*Supplement Information*). Between 1^st^ December 2021 and 16^th^ January 2022, SARS-CoV2 infection was diagnosed in 156/1121 (13.9%) patients, equating to an infection rate of 3.1 per 1000 patient days (*Supplemental Information*, **Figure S1**). Infection with Omicron was diagnosed in 145/156 (92.9%) cases; 54 (37.2%) by genotyping, 80 (55.2%) by S-gene target failure (SGTF) and 11 (7.6%) were classified as ‘probable cases’. A summary of patient characteristics of Omicron-infected and non-infected patients may be found in the *Supplemental Information*, **Table S1**. Eleven additional cases were attributed to infection by the Delta variant, with confirmation via sequencing in 9 (81.2%) cases. Of 1110 remaining patients, 71 (6.4%) were unvaccinated, 293 (26.4%) were partially vaccinated and 747 (67.3%) had received 3-doses of vaccine.

Unadjusted and adjusted vaccine effectiveness (VE) against Omicron infection was 58 (23-75)%, p=0.002, and 50 (8-71)%, p=0.018, respectively in patients who had received a booster vaccine, whilst no efficacy was seen in patients who had only been partially vaccinated (*Supplemental Information* **Table S2**). Analysing VE in the 747 patients who had been boosted, significant effectiveness was seen in both patients who received ChAdOx1, VE of 47(2-70)%, p=0.034, and BNT162b2, VE of 66 (36-81)%, p=0.0005, as their first two doses (**Figure 1**, *Supplement Information* **Table S3**).

**Figure 1.**
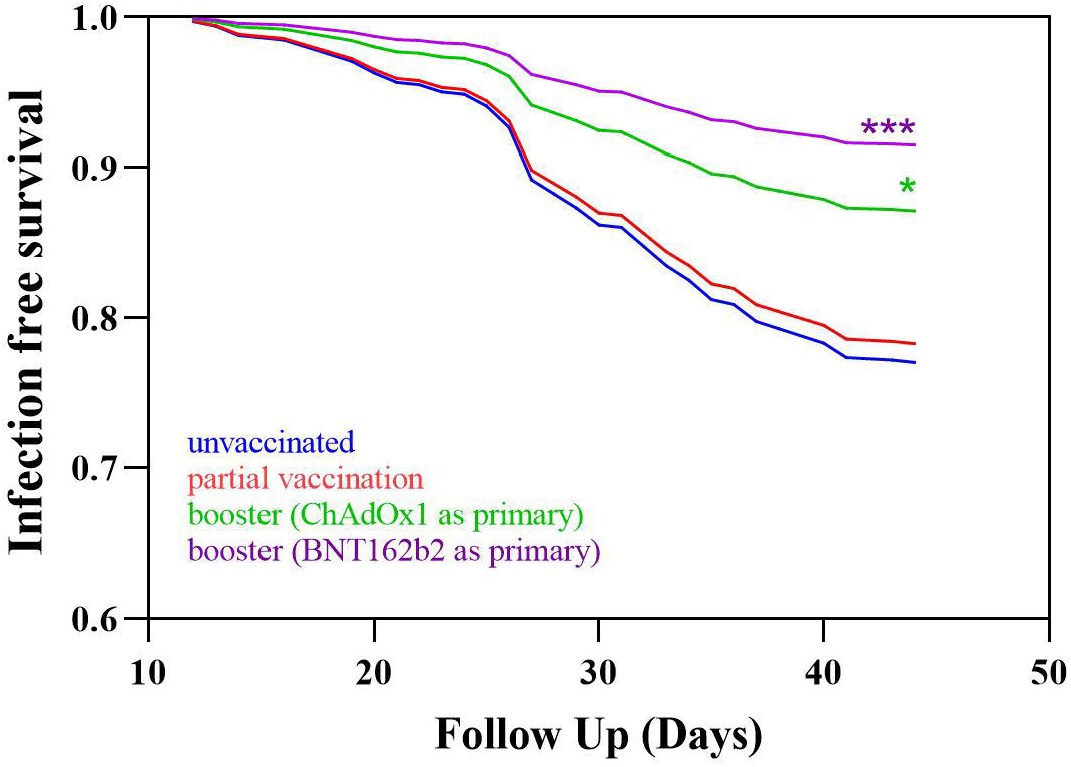
Infection event rate by vaccination status, and primary vaccine type. No difference in infection events were seen between patients who were unvaccinated and partially vaccinated, HR 0.94 (0.54-1.72), p=0.83. Patients who had received a booster dose had less infective events than unvaccinated patients whether they had received primary with ChAdOx1, HR 0.53 (0.30-0.98), p=0.034 or BNT162b2, HR 0.34 (0.19-0.64)

A total of 579/1110 (52.2%) patients had evidence of prior infection at the start of follow up, 63/145 (43.4%) patients who subsequently were diagnosed with Omicron infection and 516/965 (53.5%) patients who remained infection free. Although reinfections were seen, prior infection reduced the unadjusted and adjusted likelihood of Omicron infection, HR 0.69 (0.50-0.96), p=0.029, and HR 0.63 (0.45-0.87), p=0.0059 respectively (*Supplement Information*, **Table S2**). Analysing prior infection by vaccine status, prior infection alone, HR 0.53 (0.18-1.47), p=0.23 or prior infection with partial vaccination, HR 0.62 (0.30-1.38), p=0.20, did not reduce the likelihood of infection. For patients who were boosted, a VE of 61 (14-80)%, p=0.01 was seen in those without prior infection, and 77 (48-89)%, p=0.0001 in those with prior infection (*Supplement Information* **Table S3** and **Figure S2**).

With a median follow up of 25 (IQR: 19-28) days post diagnosis, 4/145 (2.8%) patients died within 28-days of infection. Four of 145 (2.8%) patients acquired infection via nosocomial transmission, and 2 of these patients died. Of the remaining 141 patients who were diagnosed within the outpatient setting, 12 (8.5%) were hospitalised at a median of 7 (IQR: 2.5-9.5) days post diagnosis. Seventy-six of 128 (59.4%) patients who remained outpatients received no directed therapy, compared with 5/17 (29.4%) of patients who were hospitalised either at the time, or after diagnosis (*Supplemental Information*, **Table S4**).

## Discussion

We have shown that 2-doses of a SARS-CoV2 vaccine fails to provide protection against Omicron infection. Vaccine effectiveness returns following a booster, irrespective of whether the priming was achieved with BNT162b2 or ChAdOx1. Although reinfections were common, prior infection remained clinically important in reducing the likelihood of infection, which supports immunogenicity data on breadth and durability of immune responses following infection and vaccination^7^. This may also explain why vaccination failed to demonstrate effectiveness against hospital admission, but prior infection did (*Supplemental Information*, **Table S5**). Although this also may represent a selection bias of less co-morbid patients surviving previous, more pathogenic variants.

Although immunogenicity data has shown relatively good immunological responses to SARS-CoV2 vaccines, particularly mRNA-based vaccines, in patients with ESKD on haemodialysis, responses are still weaker compared with healthy controls^6,8^. Two recent in-vitro studies have also shown the necessity of a booster dose in dialysis patients; the first showed high levels of seropositivity against the delta and omicron variants measured by spike glycoprotein cross-reactivity, whilst the second demonstrated enhanced neutralisation against Omicron following the booster dose^5,9^. The former study suggests no difference between those patients primed with ChAdOx1 compared with a mRNA vaccine, whilst the latter suggests a significant proportion of patients primed with ChAdOx1 have undetectable neutralising antibodies post 3^rd^-dose^5,9^. We saw no significant difference in clinical outcomes in this primary analysis, but this requires further monitoring.

Overall mortality rates in patients with breakthrough infection were much lower than reported in previous waves^10^. In addition to vaccination, another layer of protection against severe disease in this and other vulnerable populations was the introduction of treatments for SARS-CoV2 in non-hospitalised patients in December 2021 in the UK. Both agents available, the anti-viral Molnupiravir and neutralising antibody, Sotrovimab, have both been shown to reduce disease progression in phase 3 clinical trials^11,12^. In a common predicament however, patients with ESKD were excluded from these trials, and the effectiveness (against Omicron) and potential safety profile of these medications in haemodialysis patients is therefore unknown. Whilst no safety concerns were reported in our patient cohort, limited inference can be made on the use of molnupiravir due to numbers treated, and further assessment must be made in these patients.

In conclusion, within a haemodialysis population, 3-doses of a SARS-CoV-2 vaccine are required for clinical protection against SARS-CoV2 Omicron infection. The Omicron variant appears to result in less severe disease compared with other variants in this preliminary analysis. Despite some reassurance from this data, this multi-morbid population requires close surveillance, with rapid adaption of vaccine regimens and available treatments, as and if, evidence changes.

## Data Availability

All data produced in the present work are contained in the manuscript

## Acknowledgements

This research is supported by the National Institute for Health Research (NIHR) Biomedical Research Centre based at Imperial College Healthcare NHS Trust and Imperial College London. The authors would like to thank the West London Kidney Patient Association, all the patients and staff at ICHNT (The Imperial COVID vaccine group and dialysis staff, and staff within the North West London Pathology laboratories). The authors are also grateful for support from The Nan Diamond Fund, Sidharth and Indira Burman, and the Auchi Charitable Foundation. MP is supported by an NIHR clinical lectureship. Work in DCT’s lab is supported by a Wellcome Trust Clinical Career Development Fellowship

## References

1. Hoffmann M, Krüger N, Schulz S, et al. The Omicron variant is highly resistant against antibody-mediated neutralization: Implications for control of the COVID-19 pandemic. Cell.

2. Shuai H, Chan JF-W, Hu B, et al. Attenuated replication and pathogenicity of SARS-CoV-2 B.1.1.529 Omicron. Nature 2022.

3. Halfmann PJ, Iida S, Iwatsuki-Horimoto K, et al. SARS-CoV-2 Omicron virus causes attenuated disease in mice and hamsters. Nature 2022.

4. UK Health Security Agency. SARS-CoV-2 variants of concern and variants under investigation in England. Technical briefing 34. https://assets.publishing.service.gov.uk/government/uploads/system/uploads/attachment_data/file/1048395/technical-briefing-34-14-january-2022.pdf.

5. Carr EJ, Wu M, Harvey R, et al. Omicron neutralising antibodies after COVID-19 vaccination in haemodialysis patients. The Lancet.

6. Carr EJ, Wu M, Harvey R, et al. Neutralising antibodies after COVID-19 vaccination in UK haemodialysis patients. Lancet 2021; 398(10305): 1038–41.

7. Walls AC, Sprouse KR, Bowen JE, et al. SARS-CoV-2 breakthrough infections elicit potent, broad and durable neutralizing antibody responses. Cell.

8. Clarke CL, Martin P, Gleeson S, et al. Comparison of immunogenicity between BNT162b2 and ChAdOx1 SARS-CoV-2 vaccines in a large haemodialysis population. medRxiv 2021: 2021.07.09.21260089.

9. Faustini S, Shields A, Banham G, et al. Cross reactivity of spike glycoprotein induced antibody against Delta and Omicron variants before and after third SARS-CoV-2 vaccine dose in healthy and immunocompromised individuals. Journal of Infection.

10. Collaborative: TO, Green A, Curtis H, et al. Describing the population experiencing COVID-19 vaccine breakthrough following second vaccination in England: A cohort study from OpenSAFELY. medRxiv 2021: 2021.11.08.21265380.

11. Jayk Bernal A, Gomes da Silva MM, Musungaie DB, et al. Molnupiravir for Oral Treatment of Covid-19 in Nonhospitalized Patients. N Engl J Med 2021.

12. Gupta A, Gonzalez-Rojas Y, Juarez E, et al. Early Treatment for Covid-19 with SARS-CoV-2 Neutralizing Antibody Sotrovimab. N Engl J Med 2021; 385(21): 1941–50.

