## Supplemental Information for "Comparison of vaccine effectiveness against the Omicron (B.1.1.529) variant in patients receiving haemodialysis"

### **Methods**

***Patient Selection***

One-thousand, one-hundred and twenty-one patients who were receiving haemodialysis within Imperial College Renal and Transplant Centre on the 1st December 2021, and were being prospectively studied as part of ‘The Impact of COVID-19 on Patients with Renal disease and Immunosuppressed Patients’ study were included. The study was approved by the Health Research Authority, Research Ethics Committee (Reference: 20/WA/0123). All, infection events were captured until 16^th^ January 2022, with clinical follow up until 23^rd^ January 2022. Clinical and vaccine data were obtained from electronic patient records and the institutional vaccine database, respectively.

*Detection of SARS-CoV-2 infection*

All patients underwent weekly screening for asymptomatic infection by nasopharyngeal swabbing, with viral detection via reverse-transcriptase polymerase chain reaction (RT-PCR) assays. In addition, patients underwent additional screening for infection if symptomatic. The variant of interest, Omicron (B.1,1,529), was either detected by sequencing or latterly via the demonstration of S-gene drop out; infection was deemed of ‘probable Omicron infection’ if these criteria were not met, but overall percentage of sequenced cases within London was >96%. Reverse-transcriptase PCR was carried out using either the Thermofisher, Cepheid or Roche assays.

*SARS-CoV-2 antibody detection*

Routine serological screening of haemodialysis patients every 3-months started in June 2020, as previously described. Serum was tested for antibodies to both the nucleocapsid protein (anti-NP) and spike protein (anti-S). Anti-NP was tested using the Abbott Architect SARS-CoV-2 IgG 2 step chemiluminescent immunoassay (CMIA) according to manufacturer’s instructions. This is a non-quantitative assay and samples were interpreted as positive or negative with a threshold index value of 1.4. The presence of anti-NP was used as a marker of natural infection. For vaccine responses, anti-S IgG were assessed using the Abbott Architect SARS-CoV-2 IgG Quant II CMIA. Anti-S antibody titres are quantitative with a threshold value of 7.1 BAU/ml for positivity, and an upper level of detection of 5680 BAU/ml. Prior to December 2020, patients were initially screened for anti-NP, and those with a subthreshold anti-NP index value (0.25-1.4), underwent confirmatory testing for natural infection by assessing for receptor binding domain (anti-RBD) antibodies. This was performed using an in-house double binding antigen ELISA (Imperial Hybrid DABA; Imperial College London, London, UK), which detects total RBD antibodies

*Definition of prior SARS-CoV-2 infection*

Prior exposure was defined by a history of infection confirmed through viral detection from nasopharyngeal swab specimens, via reverse-transcriptase polymerase chain reaction (RT-PCR) assays, or by serological assessment.

***Statistical Analysis***

Statistical analysis was conducted using Prism 9.0 (GraphPad Software Inc., San Diego, California). Unless otherwise stated, all data are reported as median with interquartile range (IQR). The Chi-squared test was used for proportional assessments. The Mann-Whitney and Kruskal-Wallis tests were used to assess the difference between 2 or >2 groups, with Dunn’s post-hoc test to compare individual groups.

*Assessment of vaccine effectiveness*

Reported outcomes included RT-PCR proven SARS-CoV-2 infection, hospitalisation, and death; death was recorded as SARS-CoV2 related if it occurred within 28 days of confirmed infection. Patients who died of non-SARS-CoV2 related causes, and those who received a transplant during follow up, were censored at the time of death or transplant respectively. Event rate for infection were reported as incidence per 1000-patient days at risk. Cox proportional hazards models were used to determine adjusted hazard ratios (HR) for the first PCR-positive test post 14 days after last vaccine. Patients were categorised by vaccination status on the 1^st^ December, as unvaccinated, partially vaccinated (2 vaccines) or boosted (3 vaccines). Patients who received additional doses of a SAR-CoV-2 vaccine during the follow up period were censored at 14-days post inoculation. Only 6 patients received a 4th vaccine dose during the study period. Vaccine effectiveness (VE) was calculated using the formula VE= (1-adjusted HR) x100.

### **Figure 1. Flow diagram of study cohort.**

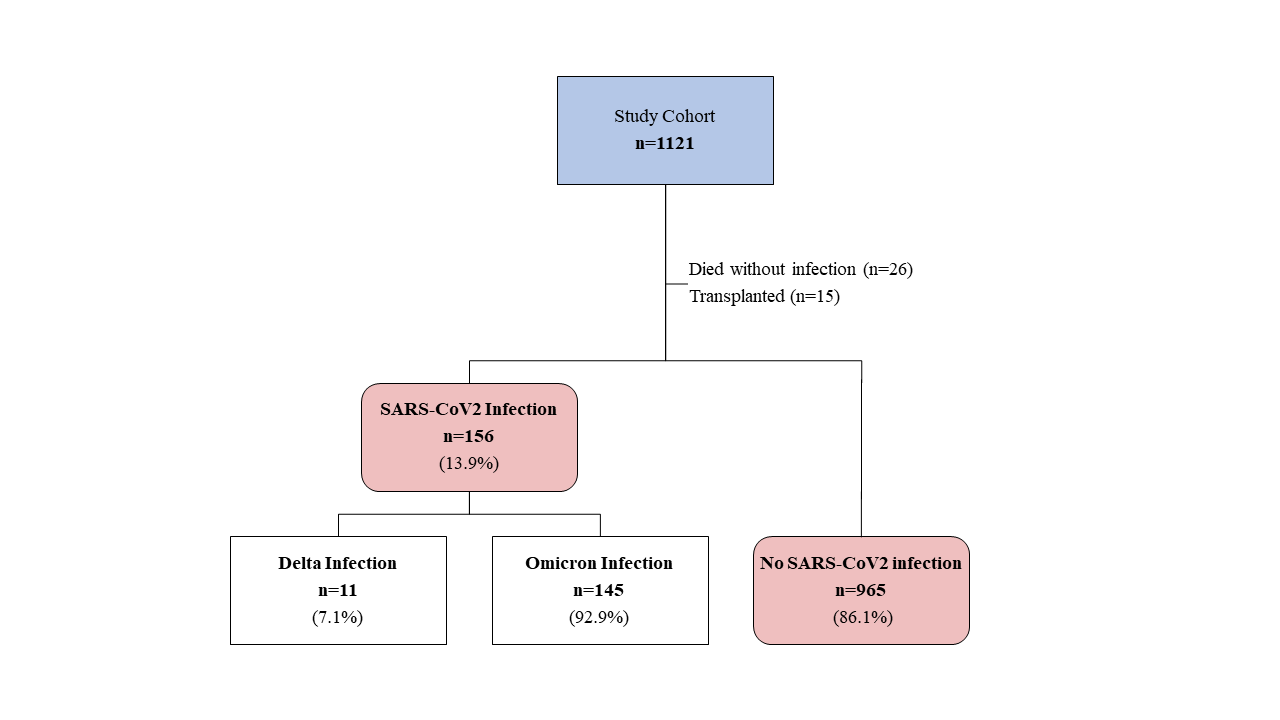

### **Table S1. Clinical characteristics associated with Omicron infection in haemodialysis patients.**

| Characteristics | | Infection Free  N=965 (%) | Omicron Infection  N= 145 (%) | p value |
| --- | --- | --- | --- | --- |
| Gender | Male  Female | 595 (61.7)  370 (38.3) | 84 (57.9)  61 (42.1) | 0.39 |
| Age | Years (Median) | 66 (56-76) | 61 (49-74) | **0.0035** |
| Ethnicity | Caucasian  Black*  Indoasian  Other | 285 (29.5)  220 (22.8)  340 (35.2)  120 (12.4) | 32 (22.1)  52 (35.9)  41 (28.3)  20 (13.8) | **0.0007** |
| Cause of ESKD | Polycystic kidney disease  Glomerulonephritis*  Diabetic nephropathy  Urological  Unknown  Other | 47 (4.9)  170 (17.6)  399 (41.3)  70 (7.3)  187 (19.4)  92 (9.5) | 4 (2.8)  20 (13.8)  65 (44.8)  6 (4.1)  39 (26.9)  11 (7.6) | 0.25 |
| Previous transplant | Yes  No | 190 (19.7)  775 (80.3) | 24 (16.6)  121 (83.4) | 0.37 |
| Immunosuppression at time of vaccine | None  Yes | 806 (83.5)  159 (16.5) | 123 (84.8)  22 (15.2) | 0.69 |
| Diabetes | No  Yes | 457 (47.4)  508 (52.6) | 71 (49.0)  74 (51.0) | 0.72 |
| Prior SARS-CoV2 infection | No  Yes | 449 (46.5)  516 (53.5) | 82 (56.6)  63 (43.4) | **0.024** |
| Vaccination status | Unvaccinated  Partially vaccinated  Boosted | 55 (5.7)  239 (24.8)  671 (69.5) | 15 (10.3)  54 (37.2)  76 (52.4) | **0.0002** |
| Vaccine type (2-doses)^ | BNT1262b2  ChAdOx1 | 495 (54.4)  415 (45.6) | 56 (43.1)  74 (56.9) | **0.0156** |

*Comparator. ^Vaccinated patients only

### **Table S2. Clinical characteristics associated with Omicron infection**

| **Variable** | **Reference Group** | **Unadjusted** | | **Adjusted** | |
| --- | --- | --- | --- | --- | --- |
|  |  | **HR (95% CI)** | **P value** | **HR (95% CI)** | **P value** |
| Age | <65 years  ≥65 years | 1  0.63 (0.45-0.88) | 0.007 | 0.74 (0.52-1.03) | 0.075 |
| Ethnicity | Black  Non-Black | 1  0.54 (0.39-0.77) | 0.0004 | 1  0.63 (0.44-0.90) | 0.01 |
| Immunosuppression | No  Yes | 1  0.94 (0.58-1.45) | 0.78 | - |  |
| Number of vaccines | Unvaccinated  Partially vaccinated  Boosted | 1  0.94 (0.54-1.72)  0.42 (0.25-0.77) | 0.83  0.0024 | 1  1.03 (0.60-1.91)  0.50 (0.29-0.92) | 0.91  0.0179 |
| Prior infection | No  Yes | 1  0.69 (0.50-0.96) | 0.0289 | 1  0.63 (0.45-0.87) | 0.0059 |

### **Table S3. Risk of SARS-CoV-2 infection by vaccination status and prior infection**

| **Variable** | **Reference Group** | **HR (95% CI)** | **P value** | **Vaccine Efficacy** |
| --- | --- | --- | --- | --- |
| **Vaccine type** | Unvaccinated  Partially vaccinated-ChAdOx1  Partially vaccinated -BNT162b2  Boosted – ChAdOx1  Boosted – BNT162b2 | 1  1.04 (0.57-1.97)  0.83 (0.43-1.62)  0.53 (0.30-0.98)  0.34 (0.19-0.64) | -  0.91  0.57  0.034  0.0005 | -  -  47 (2-70)  66 (36-81) |
| **Vaccine plus prior infection** | Unvaccinated  Unvaccinated – prior infection  Partially vaccinated  Partially vaccinated – prior infection  Boosted  Boosted – prior infection | 1  0.53 (0.18-1.47)  0.81 (0.39-1.82)  0.62 (0.30-1.38)  0.39 (0.20-0.86)  0.23 (0.11-0.52) | -  0.23  0.58  0.20  0.01  0.0001 | -  -  -  61 (14-80)  77 (48-89) |

### **Figure S2. Survival curve by vaccination status and prior infection**

There was no difference in infection events in unvaccinated patients compared unvaccinated patients with prior infection, HR 0.53 (0.18-1.47), p=0.23, partial vaccination in infection naïve patients, HR 0.81 (0.39-1.82), p=0.58 or patients with partial vaccination who were infection naïve, HR 0.62 (0.30-1.38), p=0.20. Patients who were boosted with or without prior infection experienced less Omicron infection episodes, HR 0.23 (0.11-0.52), p=0.0001 and HR 0.39 (0.20-0.86), p=0.01 respectively.

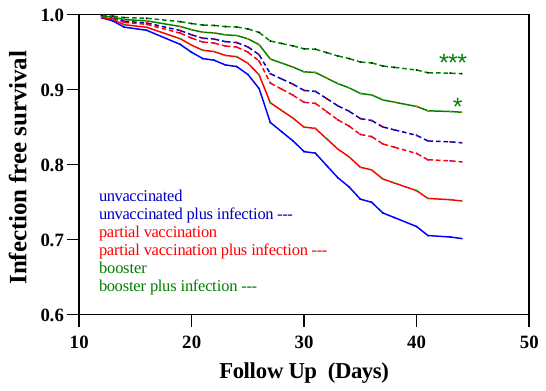

### **Table S4. Treatments received by care setting**

|  | **Care Setting** | |
| --- | --- | --- |
|  | **N=128** | **N=17*** |
| No directed therapy | 76 (59.4%) | 5 (29.4)^#^ |
| Sotrovimab | 46 (35.9%) | 2 (11.8) |
| Molnupiravir | 6 (4.7%) |  |
| Dexamethasone |  | 3 (17.6)^#^ |
| Remdesivir |  | 1 (5.9) |
| Dexamethasone plus Remdesivir |  | 2 (11.8) |
| Sotrovimab plus Remdesivir |  | 2 (11.8)^#^ |
| Dexamethasone plus Remdesivir plus Sotrovimab |  | 2 (11.8)^#^ |

*Including patients with nosocomial infection; ^#^Patients who died

### **Table S5. Vaccine effectiveness against hospitalisation by vaccination status and prior infection**

| **Variable** | **Reference Group** | **HR (95% CI)** | **P value** | **Vaccine Effectiveness** |
| --- | --- | --- | --- | --- |
| **Vaccination status** | Unvaccinated  Partially vaccinated  Boosted | 1  0.23 (0.03-1.91)  0.40 (0.10-2.63) | -  **-**  **-** |  |
| **Prior infection** | No  Yes | 1  0.27 (0.06-0.89) | -  0.049 | 73 (11-94) |
